# Detection of Antibody Responses against SARS-CoV-2 in Plasma and Saliva from Vaccinated and Infected Individuals

**DOI:** 10.1101/2021.05.11.21256972

**Authors:** Jéromine Klingler, Gregory S. Lambert, Vincenza Itri, Sean Liu, Juan C. Bandres, Gospel Enyindah-Asonye, Xiaomei Liu, Viviana Simon, Charles R. Gleason, Giulio Kleiner, Hsin-Ping Chiu, Chuan-Tien Hung, Shreyas Kowdle, Fatima Amanat, Benhur Lee, Susan Zolla-Pazner, Chitra Upadhyay, Catarina E. Hioe

## Abstract

Antibodies (Abs) are essential for the host immune response against SARS-CoV-2, and all the vaccines developed so far have been designed to induce Abs targeting the SARS-CoV-2 spike. Many studies have examined Ab responses in the blood from vaccinated and infected individuals. However, since SARS-CoV-2 is a respiratory virus, it is also critical to understand the mucosal Ab responses at the sites of initial virus exposure. Here, we examined plasma versus saliva Ab responses in vaccinated and convalescent patients. Although saliva levels were significantly lower, a strong correlation was observed between plasma and saliva total Ig levels against all SARS-CoV-2 antigens tested. Virus-specific IgG1 responses predominated in both saliva and plasma, while a lower prevalence of IgM and IgA1 Abs was observed in saliva. Antiviral activities of plasma Abs were also studied. Neutralization titers against the initial WA1 (D614G), B.1.1.7 (alpha) and B.1.617.2 (delta) strains were similar but lower against the B.1.351 (beta) strain. Spike-specific antibody-dependent cellular phagocytosis (ADCP) activities were also detected and the levels correlated with spike-binding Ig titers. Interestingly, while neutralization and ADCP potencies of vaccinated and convalescent groups were comparable, enhanced complement deposition to spike-specific Abs was noted in vaccinated versus convalescent groups and corresponded with higher levels of IgG1 plus IgG3 among the vaccinated individuals. Altogether, this study demonstrates the detection of Ab responses after vaccination or infection in plasma and saliva that correlate significantly, although Ig isotypic differences were noted. The induced plasma Abs displayed Fab-mediated and Fc-dependent functions with comparable neutralization and ADCP potencies, but a greater capacity to activate complement was elicited upon vaccination.

## Introduction

Antibodies (Abs) are an essential component of the immune responses against coronavirus disease-2019 (COVID-19). In the USA, three COVID-19 vaccines have received an authorization for emergency use from the FDA: two messenger RNA (mRNA) vaccines from Pfizer-BioNTech (BNT162b2) and Moderna (mRNA-1273), and one adenovirus-vectored vaccine from Johnson & Johnson/Janssen (Ad26.CoV2.S). All three vaccines are designed to induce Abs targeting SARS-CoV-2 spike (1,2), a membrane-anchored protein on the viral surface that contains the receptor-binding domain (RBD) necessary for binding and entry into the host cells (3–5). Other vaccines utilized in other countries also function to generate Abs against SARS-CoV-2 spike protein (6). In addition, several monoclonal Abs targeting spike protein are under development (7), and three have been authorized for emergency use by the FDA for the treatment of mild to moderate non-hospitalized COVID-19 patients: REGEN-COV (Casirivimab with Imdevimab), Eli Lilly (Bamlanivimab and Etesevimab) and Vir Biotechnology/GlaxoSmithKline (Sotrovimab).

Many studies have evaluated Ab responses against SARS-CoV-2 elicited by infection or vaccination, but most examined Abs in the blood. Considering that SARS-CoV-2 is a respiratory virus, Abs in the mucosal sites would serve as the frontline defense against this virus; however, limited data are currently available. Similarities and differences have been noted in the distribution of Ig isotypes in the blood and mucosal tissues. The primary Abs found in the blood are IgG, representing ∼75% of serum Ig. Among the four IgG subtypes, IgG1 and IgG2 comprise 66% and 23% of IgG, whereas IgG3 and IgG4 are minor components (<10% each). IgM and IgA are also abundant in blood and constitute 10% to 15% of serum Ig. IgA is the major Ab isotype of the mucosal immune system and exists as IgA1 and IgA2 (8). Of these two subtypes, IgA1 Abs predominate in both serum and secretions, but IgA2 percentages are higher in secretions than in serum. Consistent with this information, our previous study demonstrated that anti-spike Ab responses in convalescent plasma collected 1-2 months post-infection, were dominated by IgG1, although the levels varied tremendously among subjects (12). Variable levels of IgM and IgA1 were also detected and constituted the prominent Ig isotypes in some individuals. Other studies have shown that SARS-CoV-2-specific IgG, IgM and IgA responses could be detected in serum and saliva from COVID-19 patients, even though IgM and IgA declined more rapidly (9–11). However, the isotypes of vaccine-elicited Ab responses in mucosa have not been studied so far.

While the primary antiviral function of Abs is to neutralize virions, Abs also have non-neutralizing effector functions mediated via their Fc fragments. Virus-neutralizing activity was detected in IgG, IgM, and IgA fractions from COVID-19 convalescent plasma (12). COVID-19 vaccines also demonstrate the capacity to elicit potent neutralizing Ab responses (13–17). However, the full properties of Abs elicited by vaccination or infection are not yet known. In particular, limited data exists for Fc-mediated activities induced by vaccination which could play a role in vaccine efficacy (18). The binding of anti-spike Abs to virions, infected cells, or soluble spike proteins creates immune complexes capable of engaging Fc receptors (FcRs) or complement via the Abs’ Fc fragments (19,20). These interactions are determined by the Ig isotypes, as each isotype engages distinct FcRs and activates the complement system with varying potency (19,21). The FcR engagement triggers a cascade of intracellular signals critical for Fc-mediated activities, including Ab-dependent cellular phagocytosis (ADCP) and Ab-dependent cellular cytotoxicity (ADCC). Binding of C1q, the first component in the classical complement pathway, to Fc fragments on immune complexes activates the downstream complement cascade, resulting in the deposition of C3 and C4 degradation products. Depositions initiates the generation of C5 convertase and the assembly of the membrane-attack complex which is responsible for complement-mediated lysis. Complement-opsonized immune complexes also interact with complement receptors on leukocytes to trigger effector functions, including complement-dependent cell-mediated phagocytosis and cytotoxicity (22,23).

In this study, we assessed Ab responses elicited against different SARS-CoV-2 antigens from plasma and saliva samples collected from both vaccinated and convalescent donors using a multiplex bead assay that was developed in our previous study (12,24). Saliva was used as a model for oral and upper respiratory mucosal secretions, and both saliva and blood specimens from each donor were obtained simultaneously. We further compared spike- and RBD-specific Ig isotypes in the same pairs of plasma and saliva samples. Additionally, vaccine- and infection-induced plasma Abs were examined for virus neutralization and Fc-dependent functions that included ADCP, C1q binding and C3d deposition. This study provides evidence for distinct SARS-CoV-2-specific Ig isotypes in plasma compared to saliva and differences in complement binding activities associated with Ig isotype profiles.

## Methods

### Recombinant proteins

SARS-CoV-2 spike and RBD proteins were produced as described (25,26). S1, S2, and nucleoprotein antigens were purchased from ProSci Inc, CA (#97-087, #97-079 and #97-085, respectively).

### Human specimens

Plasma and saliva specimens were obtained from volunteers enrolled in Institutional Review Board-approved protocols at the Icahn School of Medicine at Mount Sinai (IRB#17-00060, IRB#19-01243) and the James J. Peter Veterans Affairs Medical Center (IRB#BAN-1604): RN#1, RN#4 and RV#1-5 after immunization; RP#2-5, 7, 12, 13 after infection; and four contemporaneous non-vaccinated COVID-19-negative subjects. Thirteen additional convalescent plasma samples (CVAP samples) were obtained from 134-229 days after symptom onset under the “Evaluation of the clinical significance of two COVID-19 serologic assays” project, which received ethical approval from the James J Peters Veterans Affairs Medical Center Quality Improvement committee. Post-immunization plasma were also collected from 20 participants in the longitudinal observational “Protection Associated with Rapid Immunity to SARS-CoV-2” (PARIS) study, which was approved by the Icahn School of Medicine at Mount Sinai Institutional Review Board (IRB#20-03374). The clinical data are summarized in **Supplementary Tables 1 and 2**. All participants signed written consent forms prior to sample and data collection. All participants provided permission for sample banking and sharing. All samples were heat-inactivated before use.

### Multiplex bead Ab binding assay

Measurement of total Ig and Ig isotypes to SARS-CoV-2 antigen-coupled beads was performed as described (12). The quantification was based on MFI values at the designated sample dilutions. For total Ig responses, specimens were diluted 4-fold from 1:100 to 1:6,400 or 102,400 (plasma) or 2-fold from 1:2 to 1:16 (saliva), reacted with antigen-coated beads, and treated sequentially with biotinylated anti-human total Ig antibodies and PE-streptavidin. Titration curves were plotted for each antigens tested and the end-point titers were determined. The isotyping assays were performed at one dilution (1:200 for plasma, 1:4 for saliva) using human Ig isotype or subclasses antibodies and the MFI values were shown. Complement deposition onto plasma Abs reactive with spike and RBD were measured according to (27) with modifications. For the C1q assay, beads with spike-Ab or RBD-Ab complexes were incubated with C1q Component from Human Serum (Sigma, #C1740) for 1 hour at room temperature, followed by an anti-C1q-PE antibody (Santa Cruz, #sc-53544 PE). For the C3d assay, Complement Sera Human (33.3%, Sigma, #S1764) was added to the beads for 1 hour at 37°C, followed by a biotinylated monoclonal anti-C3d antibody (Quidel, #A702). The relative levels of C1q and C3d deposition were obtained as MFI, from which titration curves were plotted and areas-under the curves (AUC) were calculated.

### Virus neutralization

Recombinant SARS-CoV-2 viruses encoding GFP and bearing SARS-CoV-2 spike proteins of the initial WA1 strain (D614G, designated WT), B.1.1.7 (alpha), B.1.351 (beta) or B.1.617.2 (delta) variants were used in neutralization assays as described (12,28,29). Virus infection in 293T-hACE2-TMPRSS2 cells initially seeded on collagen-coated 96-well plates was detected by GFP^+^ cells. At 18-22 hours post infection, GFP counts were acquired by the Celigo imaging cytometer (Nexcelom Biosciences, version 4.1.3.0). Each condition was tested in duplicate.

### ADCP

ADCP assays were performed using a reported protocol (30) with some modifications. FluoSpheres carboxylate-modified microspheres (Thermo Fisher, #F8823) were coupled with SARS-CoV-2 spike protein using the xMAP Antibody Coupling Kit (5 µg protein/∼36.4×10^9^ beads, Luminex #40-50016). Spike-conjugated microspheres were incubated with diluted plasma for 2 hours at 37°C in the dark. After washing and centrifugation (2,000 g, 10 minutes), the beads (∼3×10^8^ beads, 10 µL/well) were incubated with THP-1 cells (0.25×10^5^ cells, 200 µL/well) for 16 hours. The samples were analyzed on an Attune NxT flow cytometer (Thermo Fisher, #A24858). Data analysis was performed using FCS Express 7 Research Edition (De Novo Software).

### Statistical analysis

Statistical analyses were performed as designated in the figure legends using GraphPad Prism 8 (GraphPad Software, San Diego, CA).

## Results

### Detection of plasma and saliva Ab responses to SARS-CoV-2 antigens after vaccination and infection

Paired plasma and saliva specimens were collected from seven healthy recipients of COVID-19 mRNA vaccines and seven convalescent COVID-19 patients. Among vaccinees, two individuals received the Pfizer-BioNTech vaccine and five received the Moderna vaccine. Samples were collected 15-37 days after the second vaccine dose (**Supplemental Table 1A**). Convalescent patients presented with varying disease severity and donated samples 189-256 days post symptom onset (**Supplemental Table 1B**). In addition, samples from four COVID-19-negative non-vaccinated donors were tested in parallel and used to establish cut-off values.

Plasma and saliva samples were titrated for total Ig against SARS-CoV-2 spike, RBD, S1, S2, and nucleoprotein antigens. Bovine serum albumin (BSA) served as a negative control. Titration curves were ploted (**Supplemental Figure 1)**, and the end-point titers were calculated (**Figure 1A**). All plasma specimens from vaccinated subjects exhibited titrating amounts of Ig against spike, RBD, S1 and S2 above the cut-off levels, although S2 reactivity was notably lower. As expected, reactivity was not observed against nucleoprotein, apart from one sample that showed weak reactivity slightly above the cut-off value. On the other hand, convalescent plasma samples displayed titrating Ig against spike, RBD, S1, S2 as well as nucleoprotein, and S2 reactivity was again the weakest. The presence of nucleoprotein-specific Abs differentiated convalescent from vaccinated subjects; these antibodies were present in plasma from convalescent but not vaccinated subjects (**Figure 1A**, **Supplemental Figure 1**). Correlation analyses further indicated that the levels of Abs against spike and each of spike fragments (RBD, S1, S2) correlated well in both groups but no correlation was found between spike and nucleoprotein Ab levels (**Supplemental Figure 2**). A similar pattern of reactivity and correlation was seen with saliva samples (**Supplemental Figure 1, Supplemental Figure 2**), albeit saliva titers were about 3 log lower compared to plasma titers (**Figure 1A**).

**Figure 1.**
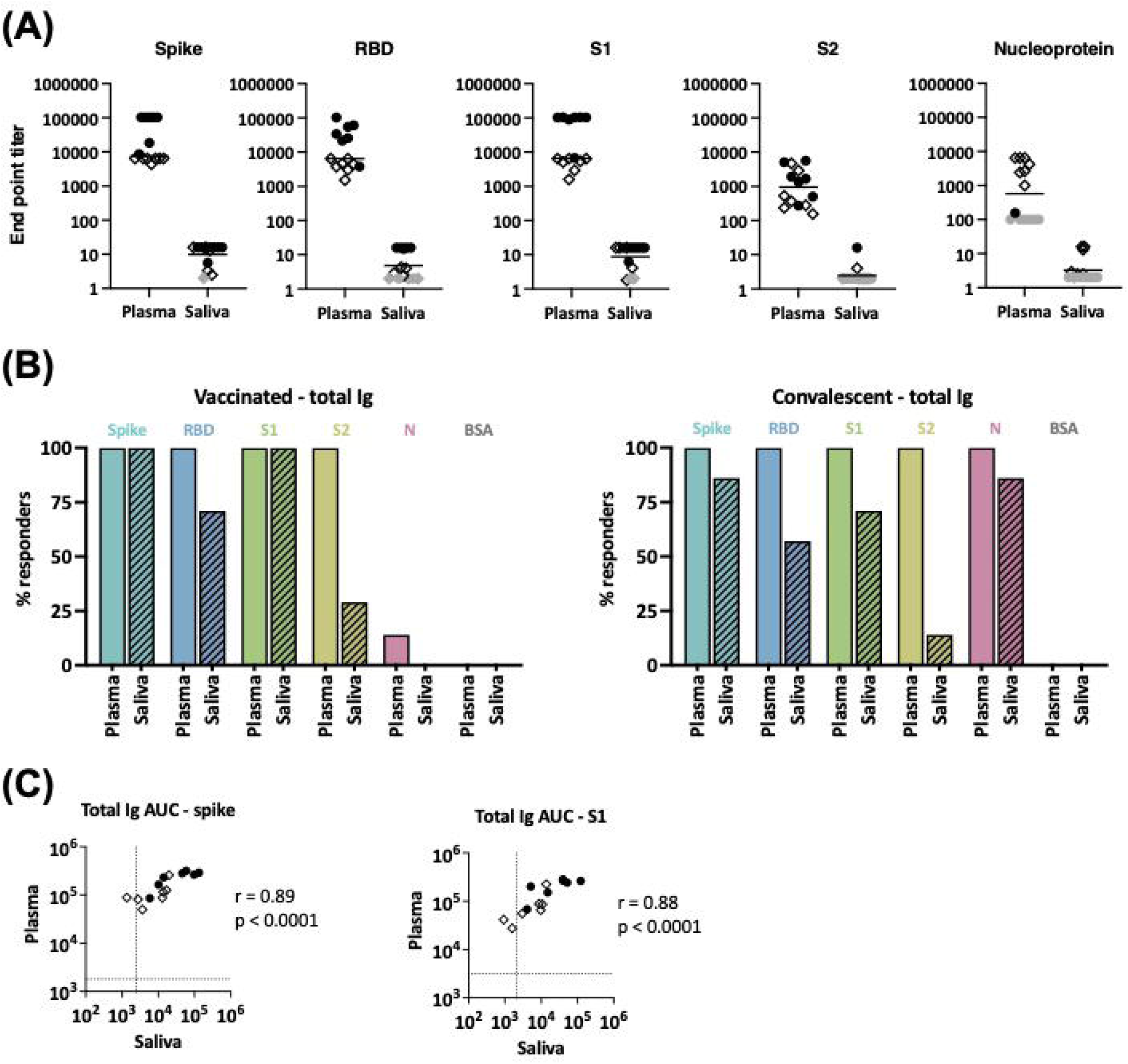
Levels of SARS-CoV-2-specific total Ig in plasma and saliva. (A) Titers of antigen-specific total Ig in plasma versus saliva specimens from vaccinated donors and convalescent COVID-19 patients. End-point titers were calculated from reciprocal dilutions that reached the the cut-off values (mean + 3SD of negative controls at the lowest dilution). Data points below the cut-off are shown at the lowest reciprocal dilutions (100 for plasma, 2 for saliva) as gray circles (vaccinated) or gray diamonds (convalescent). (B) The percentages of responders above cut-off for each antigen based on plasma versus saliva total Ig from seven vaccinated subjects (left panel) and seven convalescent COVID-19 patients (right panel). (C) Spearman correlation of spike- and S1-specific total Ig levels in plasma versus saliva from vaccinated and convalescent subjects. Areas under the curves (AUC) were calculated from the titration curves in **Supplemental Figure 1**. The dotted line indicates the cut-off value.

We then calculated the number of responders (i.e. number of individuals reaching levels above cut-offs) and found that 100% of vaccinated and convalescent subjects showed plasma Ig reactivity to spike, RBD, S1, and S2, and all convalescent subjects displayed plasma reactivity to nucleoprotein (**Figure 1B**). By contrast, only some vaccinated and convalescent subjects had positive saliva Ig reactivity. Five of the 7 saliva specimens from the vaccinated group exhibited Ig reactivity against RBD and 2 of the 7 against S2. Depending on the antigens, saliva Ig reactivity was also detected in one to six of the seven convalescent individuals. Nonetheless, the levels of total Ig in plasma and saliva correlated significantly for spike, S1, and other tested antigens (**Figure 1C and data not shown**).

### Similarities and differences in Ig isotypes against SARS-CoV-2 spike and RBD present in plasma versus saliva from vaccinated and convalescent subjects

The plasma and saliva specimens were subsequently evaluated for total Ig, IgM, IgG1-4, IgA1 and IgA2 against spike (**Figure 2A**) and RBD (**Supplemental Figure 3A**). Based on the titration data for total Ig (**Supplemental Figure 1**), plasma was tested at 1:200 dilution, while saliva was tested at 1:4 dilution. The percentage of responders for each isotype was determined using cut-off values, which were calculated as mean+3 standard deviations (SD) of the four negative specimens (**Figure 2B**).

**Figure 2.**
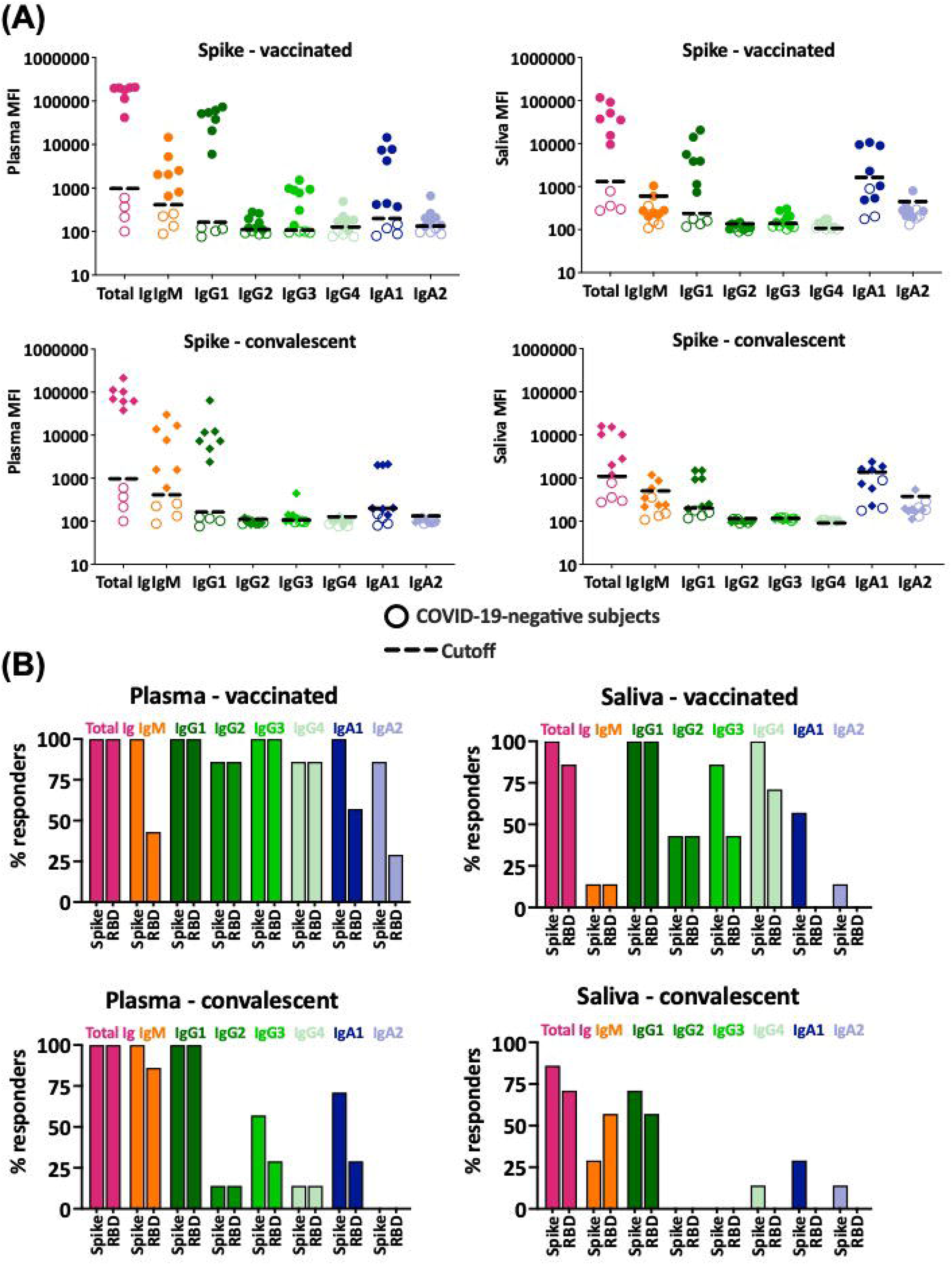
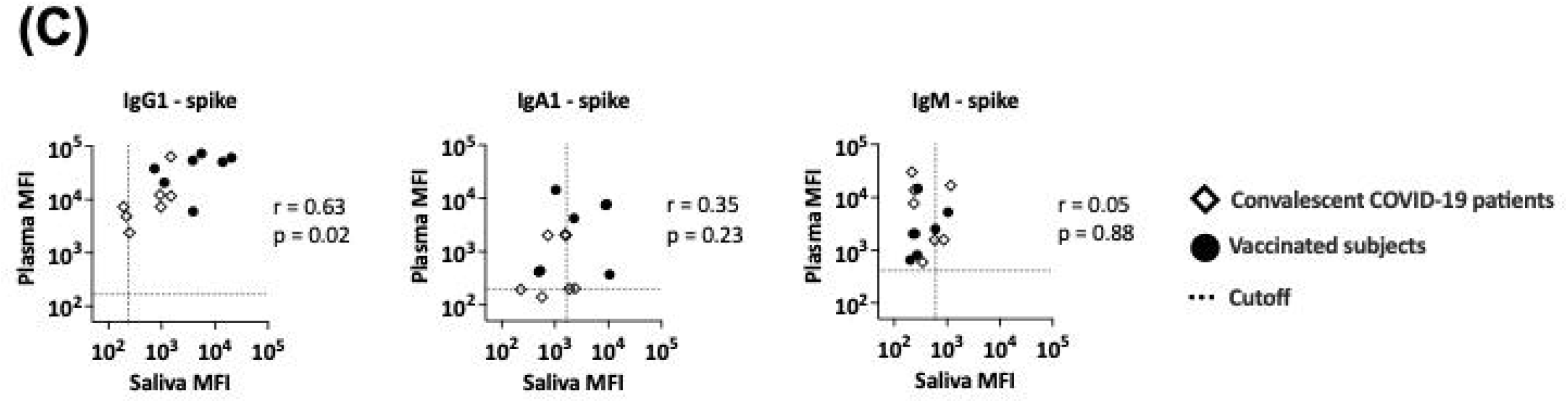
Ig isotypes against SARS-CoV-2 spike and RBD in plasma versus saliva after vaccination and infection. (A) Total Ig, IgM, IgG1, IgG2, IgG3, IgG4, IgA1 and IgA2 levels against spike were measured in plasma (left) and saliva (right) specimens from vaccinated (top panels) and convalescent COVID-19 patients (lower panels). For controls, samples from four COVID-19-negative individuals (open symbols) were tested in parallel. The dotted line represents the cut-off calculated as mean of the four control specimens + 3SD for each isotype. (B) The percentages of responders among vaccinated (top panels) and convalescent subjects (lower panels) for each spike-or RBD-specific Ig isotype on the basis of plasma (left) and saliva (right) reactivity. (C) Spearman correlation between spike- and RBD-specific IgG1, IgA1 and IgM levels in plasma versus saliva from vaccinated and convalescent subjects.

All vaccinated and convalescent plasma specimens had detectable levels of total Ig, IgM, and IgG1 against spike (**Figure 2A, B**). Similar results were observed for RBD-specific total Ig and IgG1 (**Figure 2B, Supplemental Figure 3A**), while RBD-specific plasma IgM was detected in fewer samples due to high background (**Figure 2B, Supplemental Figure 3A**), in agreement with our previous findings (12). Saliva total Ig and IgG1 against spike and RBD were also detected in most vaccinated and convalescent subjects. Interestingly, although anti-spike IgM was present in plasma from all vaccine recipients and convalescent patients, saliva IgM was detected in only a few individuals and at low levels approaching background (**Figure 2A, B**), providing evidence for the discordance in IgM responses in saliva versus plasma.

A significant proportion (>86%) of plasma specimens from vaccinated subjects displayed IgG2, IgG3, and IgG4 Abs against spike and RBD (**Figure 2B, top panels**), albeit at relatively low levels compared to IgG1 (**Figure 2A, top panels and Supplemental Figure 3**). Low levels of these minor IgG subtypes were also detected in saliva from some vaccinees (>43%). In the convalescent group, the percentages of IgG2-4 responders were much lower (**Figure 2B, bottom panels**) and the levels were near the cut-offs in plasma and saliva (**Figure 2A, bottom panels**).

Of IgA subtypes, IgA1 predominated over IgA2 in both plasma and saliva samples. Among vaccinees, 100% had plasma IgA1 Abs against spike and 86% exhibited spike-specific IgA2, although IgA2 MFI values were near the cutoff (**Figure 2A, B**). Similar results were seen for RBD-specific IgA1 and IgA2, albeit with lower percent responders and higher cut-off values (**Figure 2A, B and Supplemental Figure 3A)**. A comparable pattern was observed in convalescent plasma (**Figure 2A, B bottom panels**). The percentage of responders with specific IgA1 and IgA2 in saliva were surprisingly low. Saliva IgA1 against spike was detected in only 50% of vaccinees and 25% of convalescent patients. RBD-specific IgA1 and IgA2 against spike- and RBD were barely detected in saliva from vaccinated and convalescent subjects.

Correlation analysis of Ig isotypes in plasma versus saliva further revealed that IgG1 levels against both spike and RBD correlated strongly (**Figure 2C**). In contrast, no correlation was seen with IgM and IgA1, congruent with the differences noted in the percent IgM and IgA1 responders (**Figure 2B**). The correlation was sporadically observed for the other isotypes, but their MFI levels were near or below background (**Supplemental Figure 3B, C**).

In summary, Ab responses against spike, RBD, S1, and S2 were detected in plasma and saliva from both vaccinated and convalescent individuals, while Ab responses to nucleoprotein were detected in plasma and saliva of the convalescent group only. The dominant spike- and RBD-specific Ab isotype in saliva and plasma of both vaccinated and convalescent groups was IgG1, albeit the levels varied among individuals. IgM responses were prevalent in plasma of both vaccinated and convalescent groups but were not observed in most saliva samples. Induction of IgA1 predominated over IgA2 following vaccination and infection and was more prevalent in plasma than saliva.

### Plasma neutralizing activities against wild type versus B.1.351, B.1.1.7 and B.1.617.2 variants

Neutralizing activities by vaccine- and infection-induced plasma Abs were examined against WT and variant SARS-CoV-2 strains. Serially titrated specimens from seven vaccinated individuals, seven convalescent COVID-19, and three non-vaccinated COVID-19-negative controls were tested. Neutralization assays were performed using recombinant VSV (rVSV) expressing WT, B.1.1.7 (alpha), B.1.351 (beta), or B.1.617.2 (delta) spike proteins (29). The rVSV neutralization correlated strongly with live SARS-CoV-2 virus neutralization, demonstrating Spearman’s r >0.9 across multiple studies (29). Control samples showed background neutralization below or near 50% against all four viruses. All samples from vaccinated and convalescent groups attained >50% neutralization against WT (**Figure 3A**). In fact, all achieved near or above 90% neutralization. Similar results were observed for neutralization against B.1.1.7 and B.1.617.2. In contrast, 2 vaccinee samples and 1 convalescent specimen did not reach 50% neutralization against the B.1.351 variant. The IC_50_ titers against B.1.351 were also lower than the titers against WT (6-fold change in median) (**Figure 3B**), while the titers against B.1.1.7, B.1.617.2 and WT were not similar (**Figure 3C**). Of note, no difference was apparent in IC_50_ titers of vaccinated versus convalescent subjects against all four strains.

**Figure 3.**
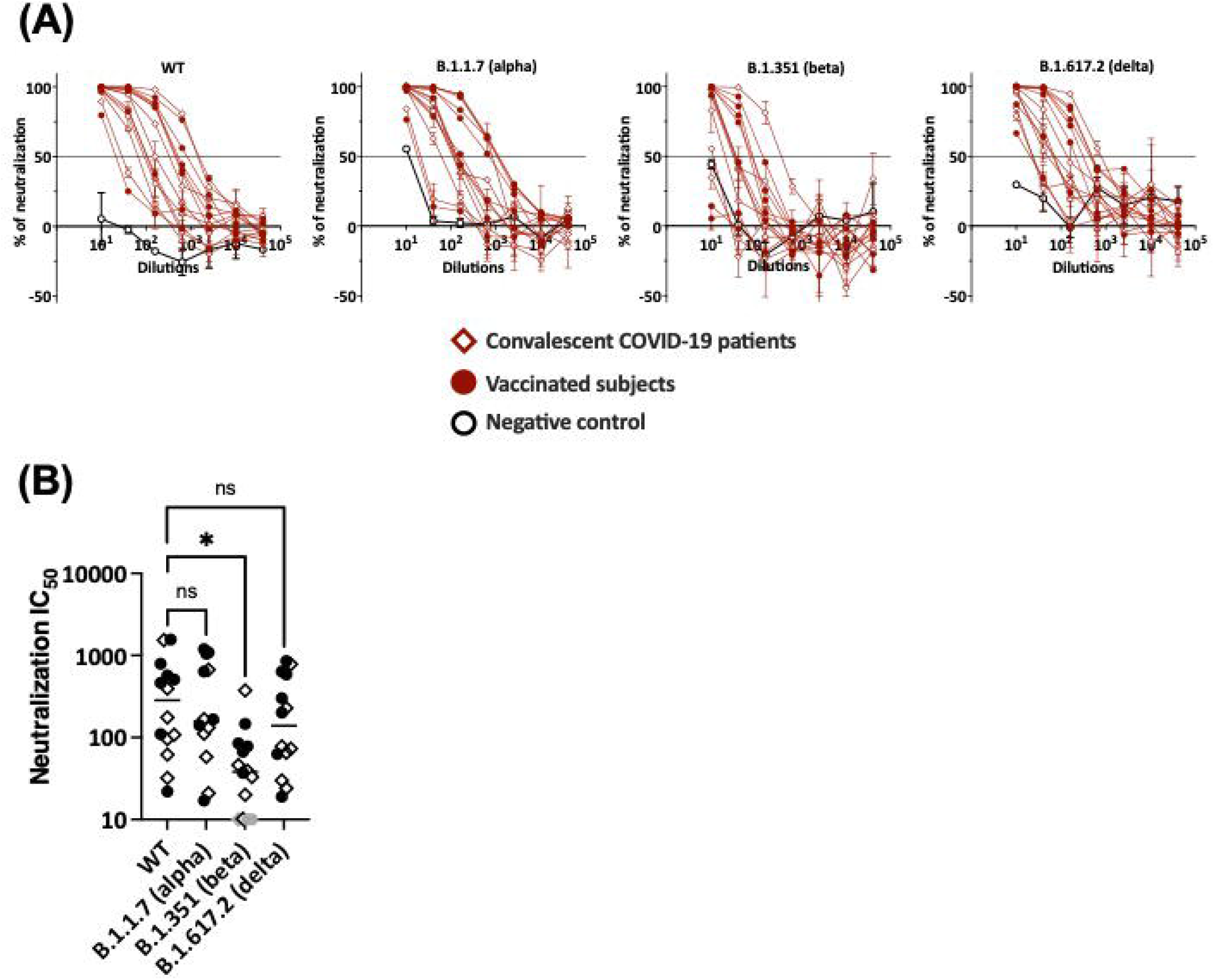
Plasma neutralization activities against WT versus variants. (A) Neutralization of recombinant VSV viruses bearing the spike proteins of SARS-CoV-2 WT, B.1.531, B.1.1.7 or B.1.617.2 by plasma specimens from vaccinated and convalescent COVID-19 donors. Plasma samples from three COVID-19-negative individuals were tested in parallel; these negative control data are shown as mean + SD of replicates from all three samples. The dotted line indicates 50% neutralization. (B) Comparison of neutralization IC50 titers against WT versus B.1.351, B.1.1.7 and B.1.617.2. The specimens that did not reach 50% neutralization were shown as gray symbols at the lowest reciprocal dilution. Statistical analysis was performed using a Kruskal-Wallis test. *: p <0.05; ns: non-significant.

### Detection of spike-specific ADCP activities in plasma of all vaccinated and convalescent donors

Because Ig isotypes are key determinants of Fc functions, we examined the Fc-mediated Ab activities from plasma specimens for both vaccinated and convalescent donors. Two Fc-dependent functions were evaluated: 1) spike-specific ADCP using THP-1 phagocytes and spike-coated fluorescent beads and 2) complement activation based on C1q and C3d deposition on spike-Ab and RBD-Ab complexes.

Spike-specific ADCP was detected above control in each of the specimens from all vaccinated and convalescent subjects (**Figure 4A, B**) and the levels corresponded with the spike-specific total Ig levels (**Figure 4C**). To assess the ADCP capacity and account for Ig level differences among samples, we calculated the AUC ratios of spike-specific ADCP over spike-binding total Ig. No difference between vaccinated and convalescent subjects was observed (**Figure 4D**). Of note, ADCP assays were performed with saliva samples, but no activity was detectable above background. When saliva was concentrated, ADCP was measurable in few specimens (data not shown), but the volumes of most samples were inadequate, precluding their assessment in this and other functional assays.

**Figure 4.**
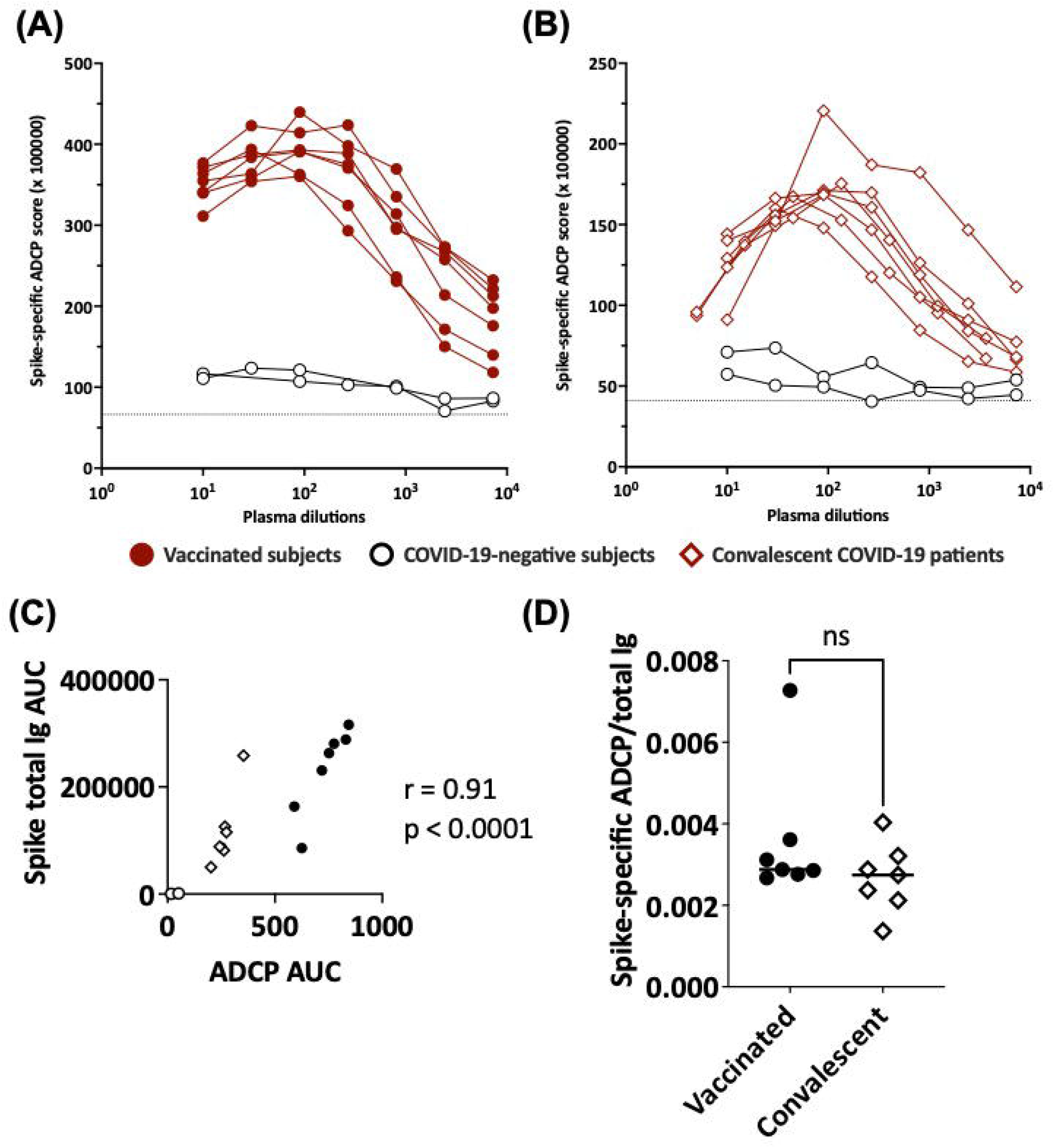
ADCP activities in plasma of vaccinated and convalescent individuals. Spike-specific ADCP activities in plasma specimens from (A) vaccinated and (B) convalescent donors were tested along with two control plasma samples from COVID-19-negative individuals. ADCP was measured by flow cytometry after incubation of plasma-treated spike-coated fluorescent beads with THP-1 phagocytes. ADCP scores were calculated as % bead^+^ cells × MFI of bead^+^ cells. The dotted line indicates the background. (C) Correlation between spike-specific ADCP AUC and spike-specific total Ig AUC from the seven vaccinated individuals, seven convalescent patients and two negative controls. (D) Ratio of spike-specific ADCP AUC to spike-specific total Ig AUC from the seven vaccinated individuals and seven convalescent patients.

### C1q and C3d deposition on spike- and RBD-specific Abs in plasma from vaccinated and convalescent subjects

C1q and C3d deposition on antigen-Ab complexes was measured utilizing antigen-coupled bead assays. The data show variability in C1q and C3d binding to spike-bound plasma Abs from both vaccinated and convalescent groups (**Figure 5A, B**). C1q and C3d binding levels correlated strongly (**Figure 5C**). Interestingly, C1q and C3d deposition was detected only in one convalescent plasma sample and at low levels (**Figure 5A, B**). In the vaccinated group, one sample did not show any C1q or C3d binding to spike, and the remaining six exhibited a range of C1q and C3d binding levels above the control. Similar results were observed with C1q and C3d binding to RBD-specific plasma Abs (**Supplemental Figure 4**). The calculated ratios of C1q or C3d AUC to total Ig AUC further indicate higher capacity of vaccine-induced Abs to bind and activate complement (**Figure 5D**). These findings were supported by data from additional 20 vaccinated and 13 convalescent donors from separate cohorts, in which the greater capacity of vaccine-induced Abs to bind C1q and C3d was even more pronounced (**Figure 5E and Supplemental Figures 5 and 6**).

**Figure 5.**
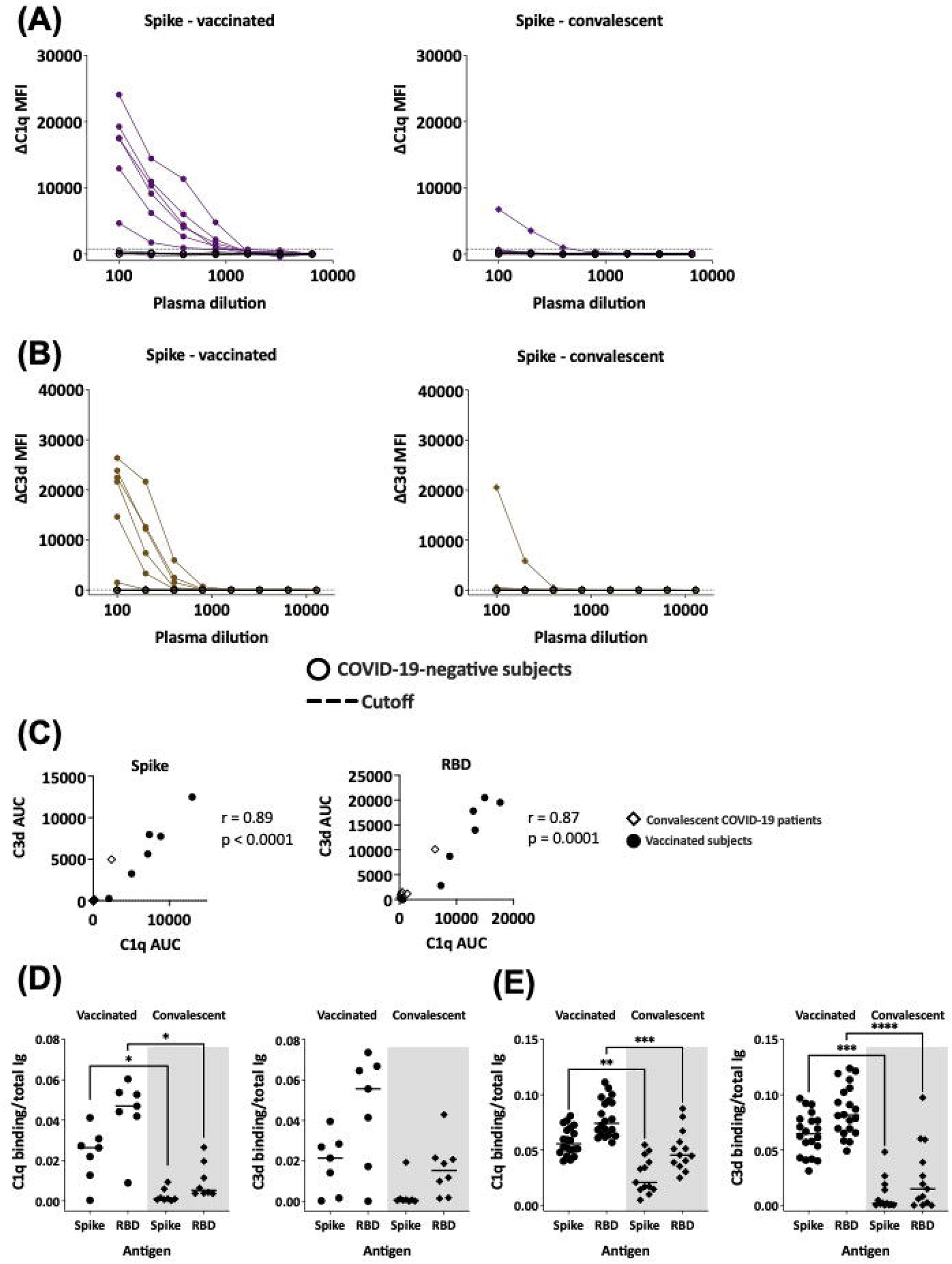
Complement-binding activities in the plasma of vaccinated and convalescent individuals. (A-B) The binding of C1q (A) and C3d (B) to spike-specific Abs in plasma specimens from vaccinated (left) and convalescent (right) donors was assessed together with four COVID-19-negative controls in multiplex bead assays. Specimens were diluted 2-fold from 1:100 to 1:6,400 or 12,800. The dotted line represents the 100x dilution cut-off values calculated as mean + 3SD of the four control specimens. ΔC1q and ΔC3d MFI values were calculated by subtracting background MFI from each assay. (C) Spearman correlation between C1q AUC and C3d AUC values for spike- or RBD-specific Abs in plasma specimens from vaccinated and convalescent donors. (D-E) Ratio of C1q and C3d binding AUC to total Ig AUC of specimens from 7 vaccinated and 7 convalescent donors (D) and from additional 20 recipients of Pfizer or Moderna mRNA vaccines and 13 convalescent donors (E). Statistical significance was assessed using a Kruskal-Wallis test (*: p<0.05, **: p<0.01, ***: p<0.001, ****: p<0.0001).

The differential complement binding activity is likely related to the relative levels of IgG subtypes generated by vaccination compared to infection. The IgG1 and IgG3 subtypes in particular have greater potency to activate the classical complement cascade (12). Indeed, the relative levels of IgG1+IgG3 over IgG2+IgG4 were higher in plasma from the vaccine group than the convalescent group (**Figure 2**). The low IgG3 levels were also observed with the larger cohort of convalescent plasma previously reported (12). To support this data, regression analyses were performed and showed that among the spike- and RBD-binding Ig isotypes tested, IgG1 and IgG3 Abs contributed most significantly to the complement binding activities (r^2^=0.74-0.95, p <0.0001).

The functional properties of plasma Abs induced after vaccination versus natural infection are summarized in **Figure 6**. The heatmap clearly shows more potent complement activation in plasma from vaccinated versus convalescent groups, even though neutralization and ADCP potencies were indistinguishable.

**Figure 6.**
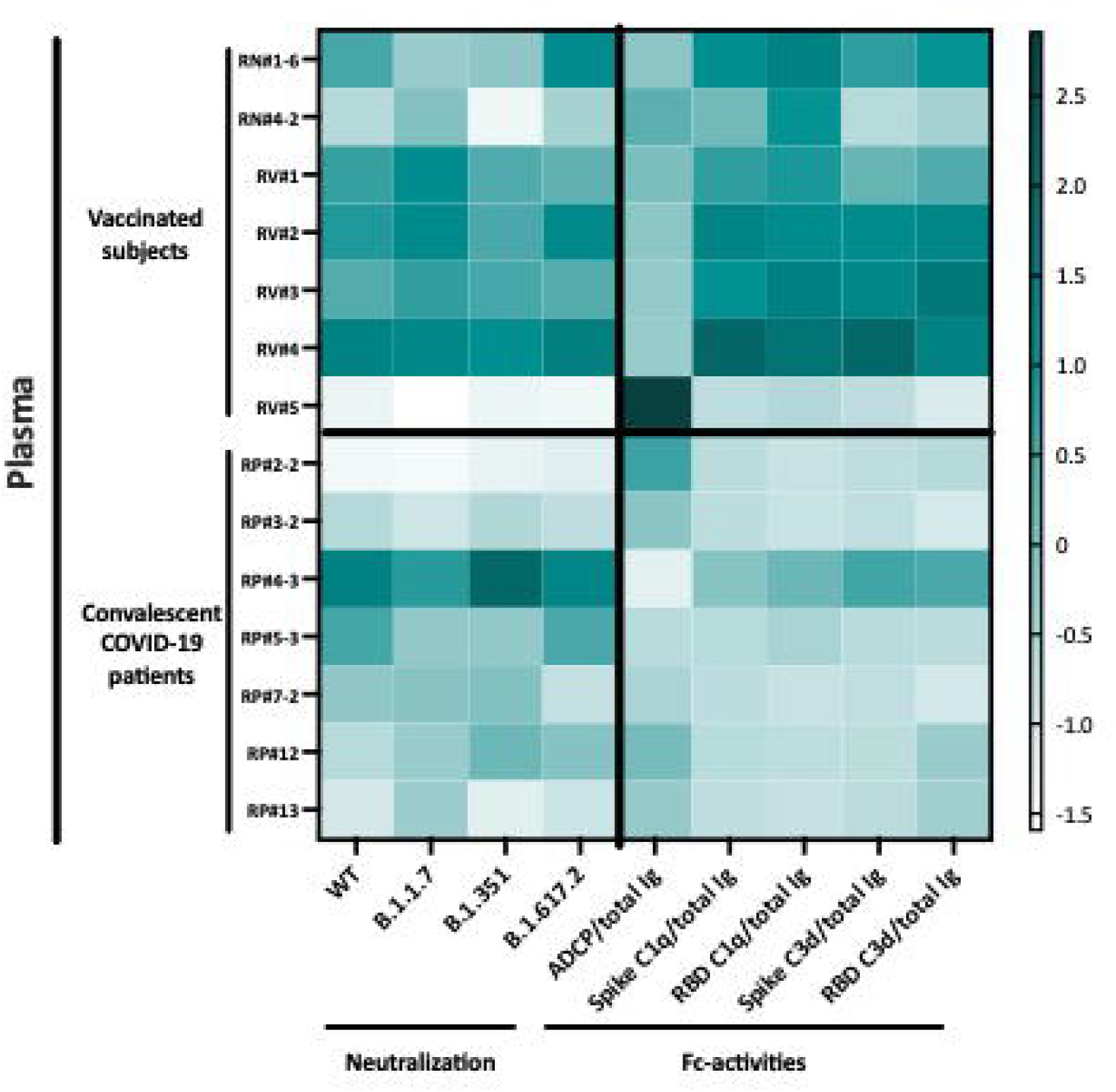
Heatmap to show the relative levels of neutralization (IC_50_) and Fc-mediated activities (ratios to total Ig) in plasma specimens from vaccinated and convalescent donors. Z-scores calculated for each Ab activity were used to generate the heatmap.

## Discussion

This study provides evidence that plasma and saliva levels of Abs elicited after vaccination or infection correlate strongly. The data bolster previous findings showing that Abs against spike and nucleoprotein were similarly detected in plasma and saliva following SARS-CoV-2 infection (31). The total levels of Abs in saliva, however, were about 100-fold lower than in plasma. Consequently, lower percentages of responders were observed for saliva versus plasma Abs, with more notable differences for S2 which induces the lowest Ab titers among the five antigens tested. Nonetheless, saliva Abs against spike, RBD, and S1 were readily detected in the majority of vaccinated and convalescent groups, and saliva Abs against nucleoprotein were detectable in all convalescent individuals tested. While these data indicate the potential use of saliva for monitoring of anti-spike Ab responses in vaccinated and convalescent individuals, lower positive responses were detected, indicating the lower sensitivity of Ab detection in saliva. Differential Ig isotypes were also seen in saliva versus plasma, although the functional implications are unclear as the antiviral activities of saliva Abs have not been investigated.

Our isotyping analysis demonstrated that IgG1 is the dominant isotype in both plasma and saliva from all vaccinated individuals and convalescent patients. However, the IgM and IgA levels were lower in saliva versus plasma. This contrasts to recent findings in milk from convalescent mothers where the dominant spike-specific Ab responses were IgA and this response was not necessarily associated with induction of IgG or IgM Abs (32). However, after vaccination milk Ab responses were dominated by IgG (33). Our data further show that compared to natural infection, vaccination induces a higher prevalence for IgG2-4 Abs both in plasma and saliva, albeit at relatively low levels. Of note, 100% of plasma from vaccinated subjects had detectable spike and RBD-specific IgG3 Abs, while only 57% and 29% responders were observed for convalescent plasma, respectively. The samples tested in this study were obtained >189 days post symptom onset, but the pattern was similar to that seen in convalescent plasma collected earlier (<8 weeks post symptom onset, 7-17% responders for spike- and RBD-specific IgG3 Abs) (12), indicating that this IgG subtype profile is maintained throughout the observation period.

We examined the potential plasma neutralization against the initial Seattle WA1 strain (WT) and SARS-CoV-2 variants of concern (B.1.1.7, B.1.351, B.1.617.2), and observed potent neutralization activity against WT in each of the studied samples. In agreement with published reports (34,35), weaker neutralization activities were seen against B.1.351 (beta), while neutralization of B1.1.7 (alpha) and B.1.617.2 (delta) was comparable to that of WT (36–39). No difference was seen in the IC_50_ titers against each of these four viruses between vaccine and convalescent groups. The neutralization titers against WT after vaccination were also similar to those of convalescent samples collected at earlier time points (<8 weeks after symptom onset) (12). The effects of these mutations on the non-neutralizing Fc-dependent functions are yet to be determined. Similar to neutralization, spike-specific ADCP activities were detected in plasma from all vaccinated and convalescent individuals. However, no correlation was observed between neutralization and ADCP activities (data not shown). Moreover, complement binding activities were distinct from neutralization and ADCP, suggesting that these functions may be mediated by distinct Ig populations or by Abs targeting different epitopes. The ability to thwart neutralization may offer an advantage to the transmissibility of these and other emerging variants, but the significance of Fc-mediated antiviral activities remains unclear.

While neutralization and ADCP capacity induced by vaccination and infection were indistinguishable, vaccine-induced plasma Abs displayed a more robust ability to mediate complement binding and activation as compared to infection-induced counterparts. The differential potencies were apparent when Ab levels were considered and when comparison was made with infection-induced Abs from earlier or later time points (data not shown). Evaluation of more plasma samples independently collected from separate cohorts of vaccinated and convalescent donors revealed a consistent pattern with significantly greater capacities of vaccine-induced Abs to bind and activate complement. The mRNA vaccine-induced Ab responses were also reported in recent work with >8000 finger stick blood specimens to have higher seroconversion rates and greater cross-reactivity with SARS-CoV-1 and Middle Eastern respiratory syndrome (MERS)-CoV RBDs (40), implying the superior quality of vaccine-induced Ab responses. Greater complement deposition activity was associated with higher levels of IgG1 and IgG3, the two IgG subtypes with the highest potency for complement fixation. Nonetheless, our study was limited by its relatively small sample sizes that are also restricted to mRNA vaccine recipients and sample collection at only one time point, precluding us from evaluating Abs elicited by other types of vaccines and from assessing changes of vaccine-induced responses over time. Analysis of longitudinally collected specimens with a larger sample size from recipients of different COVID-19 vaccines are warranted to reach definitive conclusions.

In addition to Ig isotypes, a parameter known to influence complement binding is Fc glycosylation, as the removal of terminal galactose from IgG Fc glycans has been shown to decrease C1q binding and downstream classical complement activation without affecting FcγR-mediated functions (41,42). Similarly, the sialylation of IgG Fc domains has been demonstrated to impair complement-dependent cytotoxicity (43). The Fc glycan compositions of vaccine- and infection-induced Abs are yet unknown. The importance of complement binding/activation for protection against SARS-CoV-2 also requires further investigation. It should be noted, however, that human neutralizing monoclonal Abs against SARS-CoV-2 requires a functional Fc region capable of binding complement and engaging FcγR for ADCP and ADCC, for optimal protection therapy (44).

In conclusion, this study demonstrated that saliva and plasma Ab responses against SARS-CoV-2 antigens were elicited following vaccination or infection. Ab responses in plasma and saliva correlated significantly, although Ig isotypic differences were noted between the vaccinated and convalescent individuals. Moreover, vaccination- and infection-induced plasma Abs exhibited Fab-mediated and Fc-dependent functions that included neutralization against WT and variants, phagocytosis, and complement activation. This study provide initial evidence for a superior potency of vaccine-induced Abs against spike to activate complement via the classical pathway, although the clinical significance of this function remains unclear and requires further investigation.

## Data Availability

Raw data are available upon written request to the corresponding author.

## Author contributions

J.K. and C.E.H. wrote the manuscript. J.K., G.S.L., S.Z-P, C.U., and C.E.H. designed the experiments. J.K., G.S.L., V.I., and X.L. performed the experiments and collected the data. J.K., G.S.L., B.L., S.Z-P., C.U., and C.E.H. analyzed the data. H-P.C., C-T.H., S.K., F.A., and B.L. provided protocols, antigens, cells and virus stocks. G.E-A., J.C.B., and S.L. obtained specimens. V.S., C.R.G. and G.K. provided specimens. All authors read and approved the final manuscript.

## Acknowledgments

We thank Dr. Florian Krammer for providing spike and RBD antigens, and all the donors for their contribution to the research. We would like to thank the expertise and assistance of Dr. Christopher Bare and the Dean’s Flow Cytometry CORE at Mount Sinai. We would like to thank Rozita Emami-Gorizi and Jameel Z. Iqbal at the James J. Peters VA Medical Center, the PARIS study team (Hala Alshammary, Angela A Amoako, Dalles Andre, Mahmoud H Awawda, Katherine F Beach, Maria C Bermúdez-González, Juan Manuel Carreno, Gianna Cai, Rachel L Chernet, Christian Cognigni, Karine David, Lily Q Eaker, Emily D Ferreri, Daniel L Floda, Hisaaki Kawabata, Florian Krammer, Giulio Kleiner, Neko Lyttle, Wanni A Mendez, Lubbertus C F Mulder, Ismail Nabeel, Annika Oostenink, Ariel Raskin, Aria Rooker, Kayla T Russo, Ashley Beathrese T Salimbangon, Miti Saksena, Amber S Shin, Gagandeep Singh, Viviana Simon, Levy A Sominsky, Komal Srivastava, Johnston Tcheou, Ania Wajnberg) and the Personalized Virology Initiative team at the Simon lab at Mount Sinai for the participants recruitment and sample processing.

## Supplementary figure legends

**Supplemental Figure 1**. Titration curves are shown for total Ig against spike, RBD, S1, S2, nucleoprotein (N), and BSA in plasma and saliva specimens from seven vaccinated subjects (left panels), seven convalescent COVID-19 patients (right panels) and four COVID-19-negative subjects (gray). Specimens were diluted at 4-fold dilutions from 1:100 to 1:6,400 or 102,400 (plasma) or 2-fold from 1:2 to 1:16 (saliva). The dotted lines indicated the cut-off values calculated as mean + 3SD of 1:100 diluted plasma or 1:4 diluted saliva of the four COVID-19-negative specimens. Data were generated from the multiplex bead antibody binding assay and mean fluorescent intensity (MFI) values were plotted.

**Supplemental Figure 2**. Spearman correlation of spike-versus RBD-, S1-, S2- or nucleoprotein-specific total Ig levels in (A) plasma or (B) saliva from vaccinated and convalescent subjects.

**Supplemental Figure 3**. (A) Total Ig, IgM, IgG1, IgG2, IgG3, IgG4, IgA1 and IgA2 levels against RBD were measured in plasma (left) and saliva (right) specimens from vaccinated (top panel), convalescent COVID-19 patients (lower panel) and COVID-19-negative controls. The dotted line represents the cut-off values calculated as mean of the four control specimens + 3SD for each isotype. (B-C) Spearman correlation of (B) spike- and (C) RBD-specific isotypes levels in plasma versus saliva from vaccinated and convalescent subjects. The dotted line represents the cut-off.

**Supplemental Figure 4**. (A-B) C1q or C3d binding to RBD-specific Abs in plasma specimens from seven vaccinated individuals (left) and seven convalescent COVID-19 patients (right) and four COVID-19-negative controls. Specimens were diluted at 2-fold dilutions from 1:100 to 1:6,400 or 12,800. The dotted line represents the 100x dilution cut-off calculated as mean of the four control specimens + 3SD. ΔC1q (A) and ΔC3d (B) MFI values were calculated by subtracting background MFI from each assay.

**Supplemental Figure 5**. Titration curves are shown for total Ig against spike, RBD, S1, S2 and nucleoprotein in sera and plasma specimens from 20 vaccinated subjects (left panels), 13 convalescent COVID-19 patients (right panels) and four COVID-19-negative subjects (gray). Specimens were diluted at 4-fold dilutions from 1:100 to 1:12,800. The dotted lines indicated the cut-off values calculated as mean + 3SD of 1:100 diluted plasma of the four COVID-19-negative specimens. Data were generated using the multiplex bead antibody binding assay and mean fluorescent intensity (MFI) values were plotted.

**Supplemental Figure 6**. (A-B) C1q or C3d binding to spike- or RBD-specific Abs in plasma specimens from 20 vaccinated individuals (left) and 13 convalescent COVID-19 patients (right) and four COVID-19-negative controls. Specimens were diluted at 2-fold dilutions from 1:100 to 1:6,400 or 12,800. The dotted line represents the 100x dilution cut-off calculated as mean of the four control specimens + 3SD. ΔC1q (A) and ΔC3d (B) MFI values were calculated by subtracting background MFI from each assay.

