## Supplemental Figures 1-6 for "Detection of Antibody Responses against SARS-CoV-2 in Plasma and Saliva from Vaccinated and Infected Individuals"

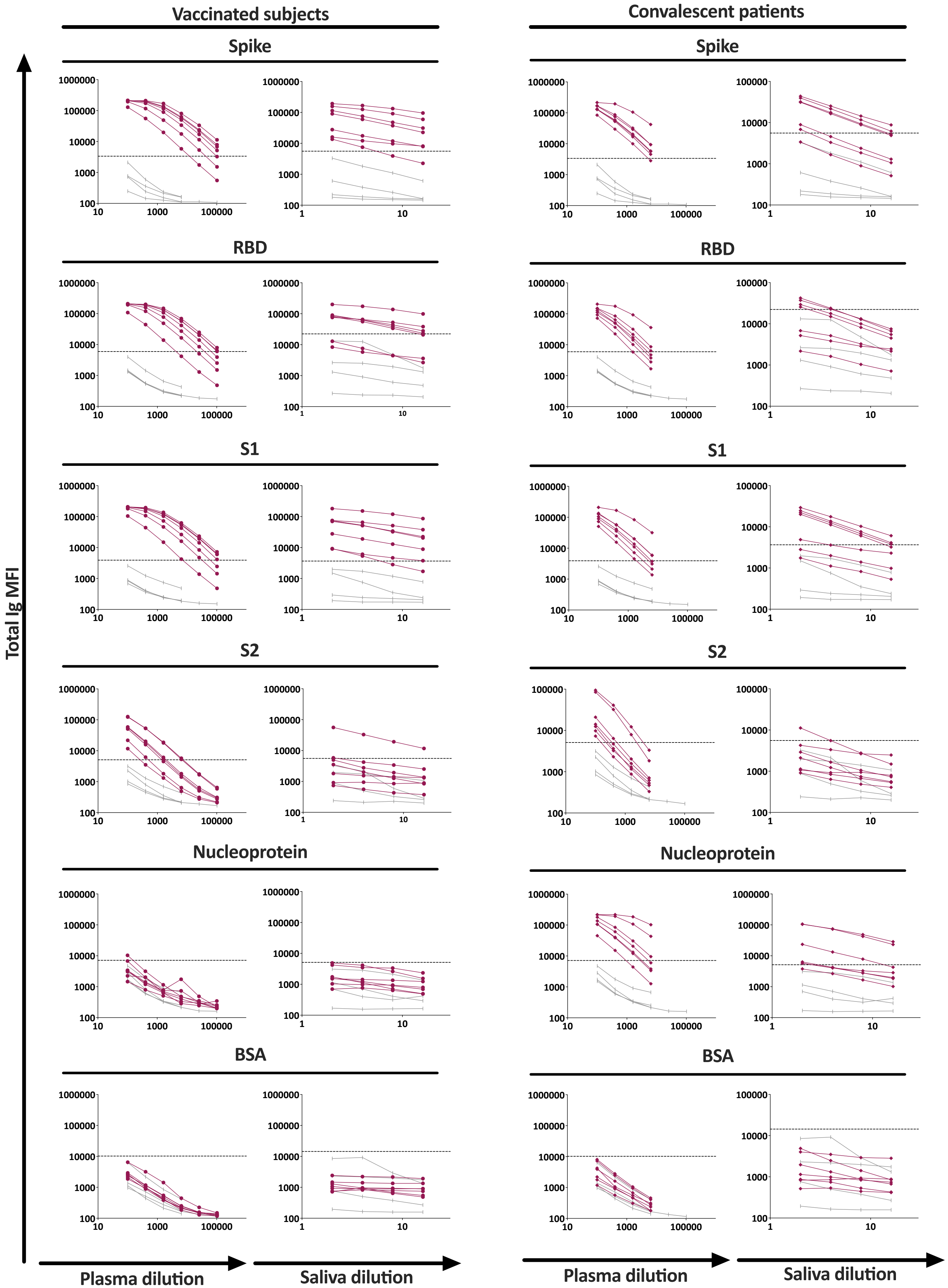

Vaccinated subjects
Convalescent patients
Spike
RBD
S1
S2
Nucleoprotein
BSA
Total Ig MFI
Plasma dilution
Saliva dilution

**Supplemental Figure 1.** Titration curves are shown for total Ig against spike, RBD, S1, S2, nucleoprotein (N), and BSA in plasma and saliva specimens from seven vaccinated subjects (left panels), seven convalescent COVID-19 patients (right panels) and four COVID-19-negative subjects (gray). Specimens were diluted at 4-fold dilutions from 1:100 to 1:6,400 or 102,400 (plasma) or 2-fold from 1:2 to 1:16 (saliva). The dotted lines indicated the cut-off values calculated as mean + 3SD of 1:100 diluted plasma or 1:4 diluted saliva of the four COVID-19-negative specimens. Data were generated from the multiplex bead antibody binding assay and mean fluorescent intensity (MFI) values were plotted.

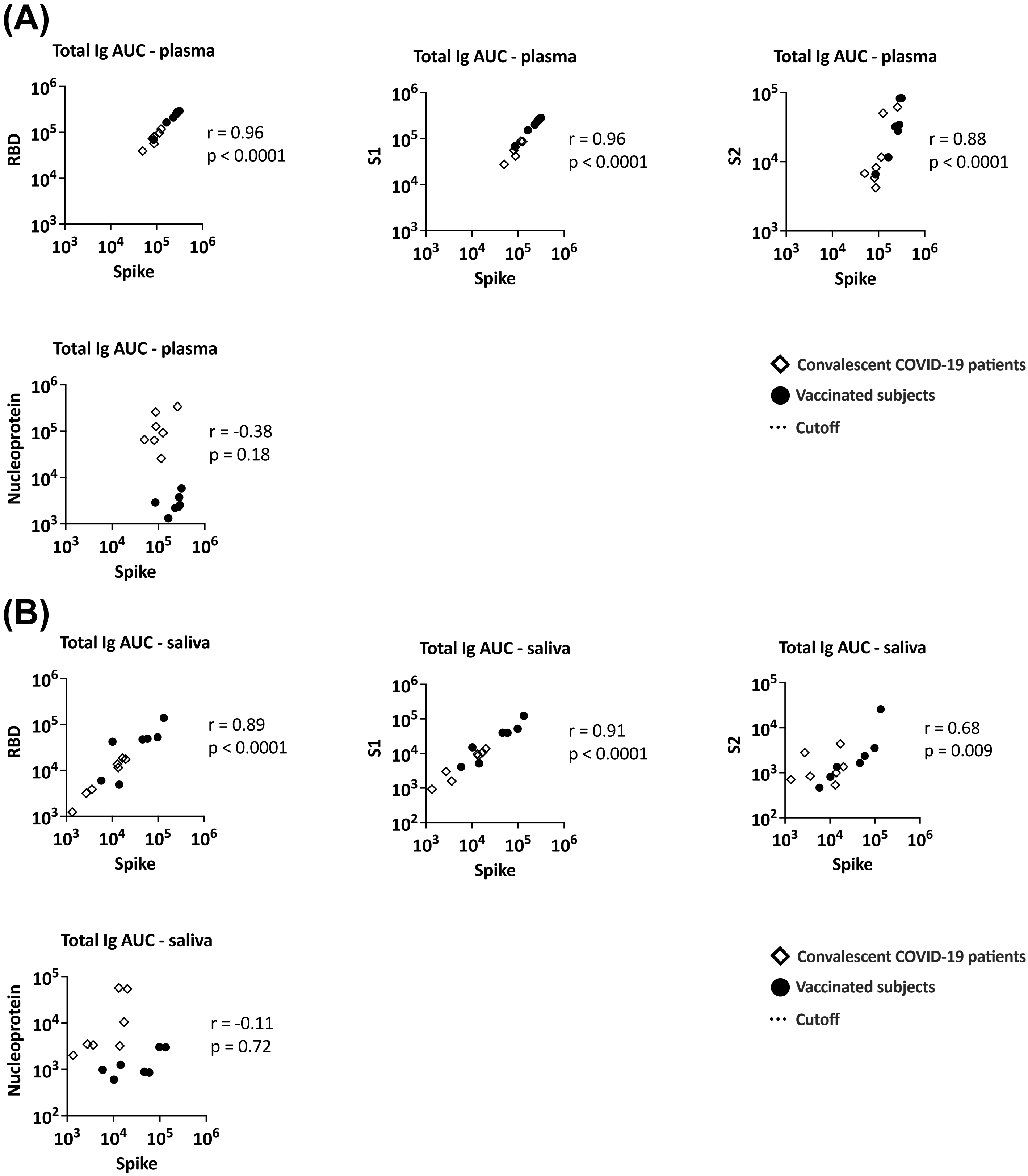
(A)
Total Ig AUC - plasma
RBD
Spike
r = 0.96
p < 0.0001
Total Ig AUC - plasma
S1
Spike
r = 0.96
p < 0.0001
Total Ig AUC - plasma
S2
Spike
r = 0.88
p < 0.0001
Total Ig AUC - plasma
Nucleoprotein
Spike
r = -0.38
p = 0.18
Convalescent COVID-19 patients
Vaccinated subjects
Cutoff
(B)
Total Ig AUC - saliva
RBD
Spike
r = 0.89
p < 0.0001
Total Ig AUC - saliva
S1
Spike
r = 0.91
p < 0.0001
Total Ig AUC - saliva
S2
Spike
r = 0.68
p = 0.009
Total Ig AUC - saliva
Nucleoprotein
Spike
r = -0.11
p = 0.72
Convalescent COVID-19 patients
Vaccinated subjects
Cutoff

**Supplemental Figure 2.** Spearman correlation of spike- versus RBD-, S1-, S2- or nucleoprotein-specific total Ig levels in (A) plasma or (B) saliva from vaccinated and convalescent subjects.

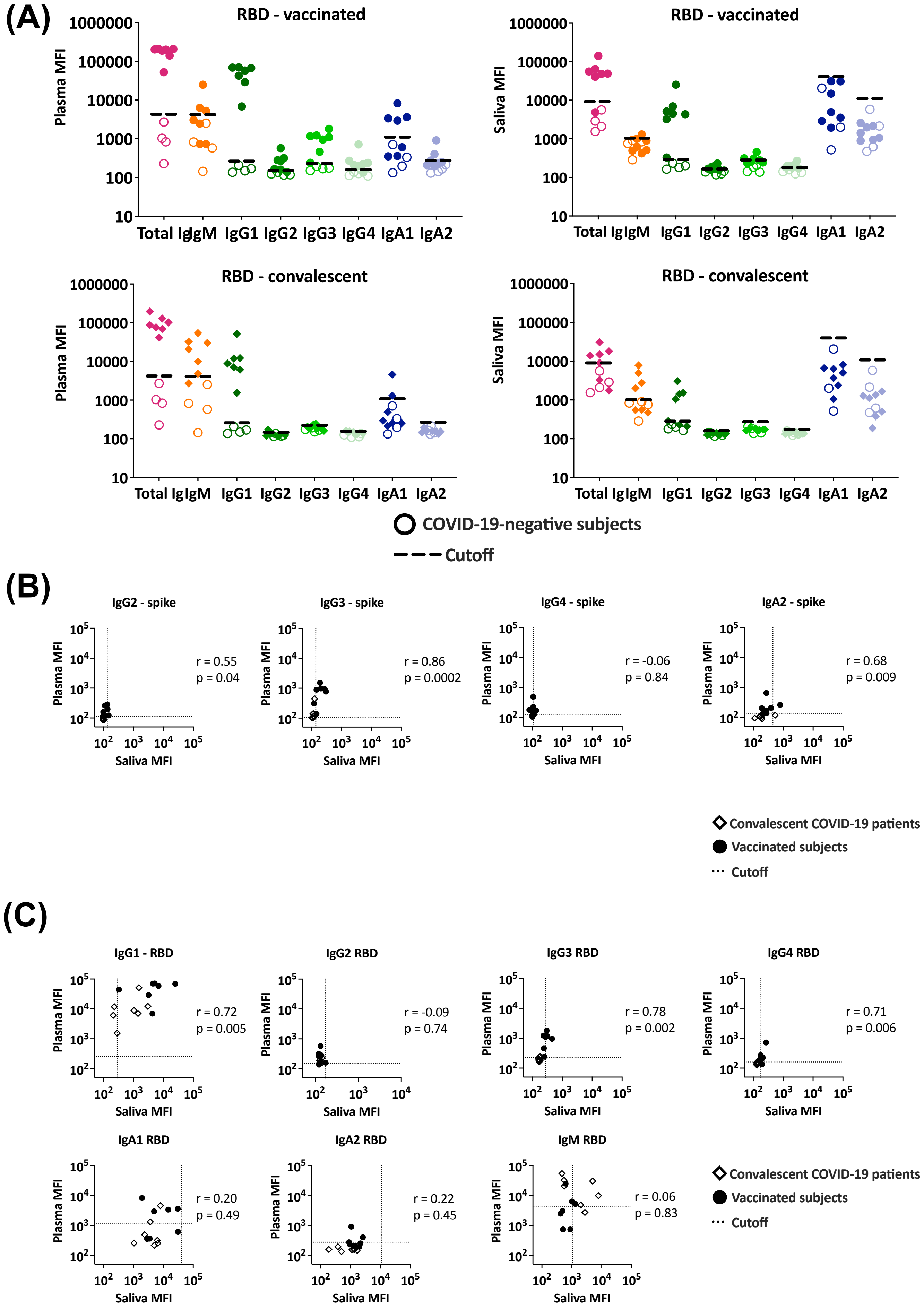
(A)
RBD - vaccinated
Plasma MFI
Saliva MFI
1000000
100000
10000
1000
100
10
Total Ig
IgM
IgG1
IgG2
IgG3
IgG4
IgA1
IgA2
RBD - convalescent
COVID-19-negative subjects
Cutoff
(B)
IgG2 - spike
r = 0.55
p = 0.04
IgG3 - spike
r = 0.86
p = 0.0002
IgG4 - spike
r = -0.06
p = 0.84
IgA2 - spike
r = 0.68
p = 0.009
Saliva MFI
Plasma MFI
Convalescent COVID-19 patients
Vaccinated subjects
Cutoff
(C)
IgG1 - RBD
r = 0.72
p = 0.005
IgG2 RBD
r = -0.09
p = 0.74
IgG3 RBD
r = 0.78
p = 0.002
IgG4 RBD
r = 0.71
p = 0.006
IgA1 RBD
r = 0.20
p = 0.49
IgA2 RBD
r = 0.22
p = 0.45
IgM RBD
r = 0.06
p = 0.83
Saliva MFI
Plasma MFI
Convalescent COVID-19 patients
Vaccinated subjects
Cutoff

**Supplemental Figure 3.** (A) Total Ig, IgM, IgG1, IgG2, IgG3, IgG4, IgA1 and IgA2 levels against RBD were measured in plasma (left) and saliva (right) specimens from vaccinated (top panel), convalescent COVID-19 patients (lower panel) and COVID-19-negative controls. The dotted line represents the cut-off values calculated as mean of the four control specimens + 3SD for each isotype. (B-C) Spearman correlation of (B) spike- and (C) RBD-specific isotypes levels in plasma versus saliva from vaccinated and convalescent subjects. The dotted line represents the cut-off.

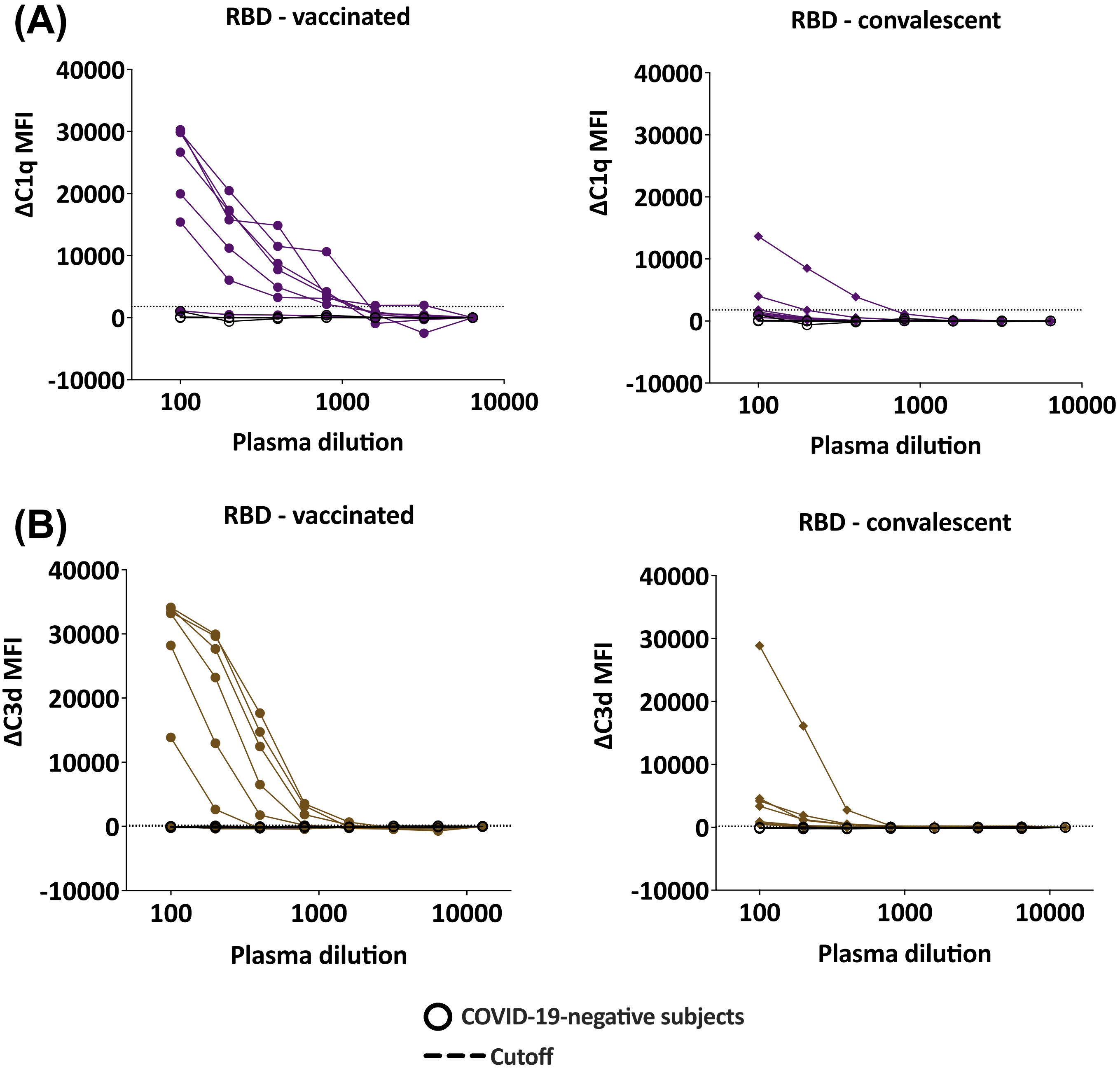
(A)
RBD - vaccinated
ΔC1q MFI
40000
30000
20000
10000
0
-10000
100
1000
10000
Plasma dilution
RBD - convalescent
ΔC1q MFI
40000
30000
20000
10000
0
-10000
100
1000
10000
Plasma dilution
(B)
RBD - vaccinated
ΔC3d MFI
40000
30000
20000
10000
0
-10000
100
1000
10000
Plasma dilution
RBD - convalescent
ΔC3d MFI
40000
30000
20000
10000
0
-10000
100
1000
10000
Plasma dilution
COVID-19-negative subjects
Cutoff

**Supplemental Figure 4.** (A-B) C1q or C3d binding to RBD-specific Abs in plasma specimens from seven vaccinated individuals (left) and seven convalescent COVID-19 patients (right) and four COVID-19-negative controls. Specimens were diluted at 2-fold dilutions from 1:100 to 1:6,400 or 12,800. The dotted line represents the 100x dilution cut-off calculated as mean of the four control specimens + 3SD. ΔC1q (A) and ΔC3d (B) MFI values were calculated by subtracting background MFI from each assay.

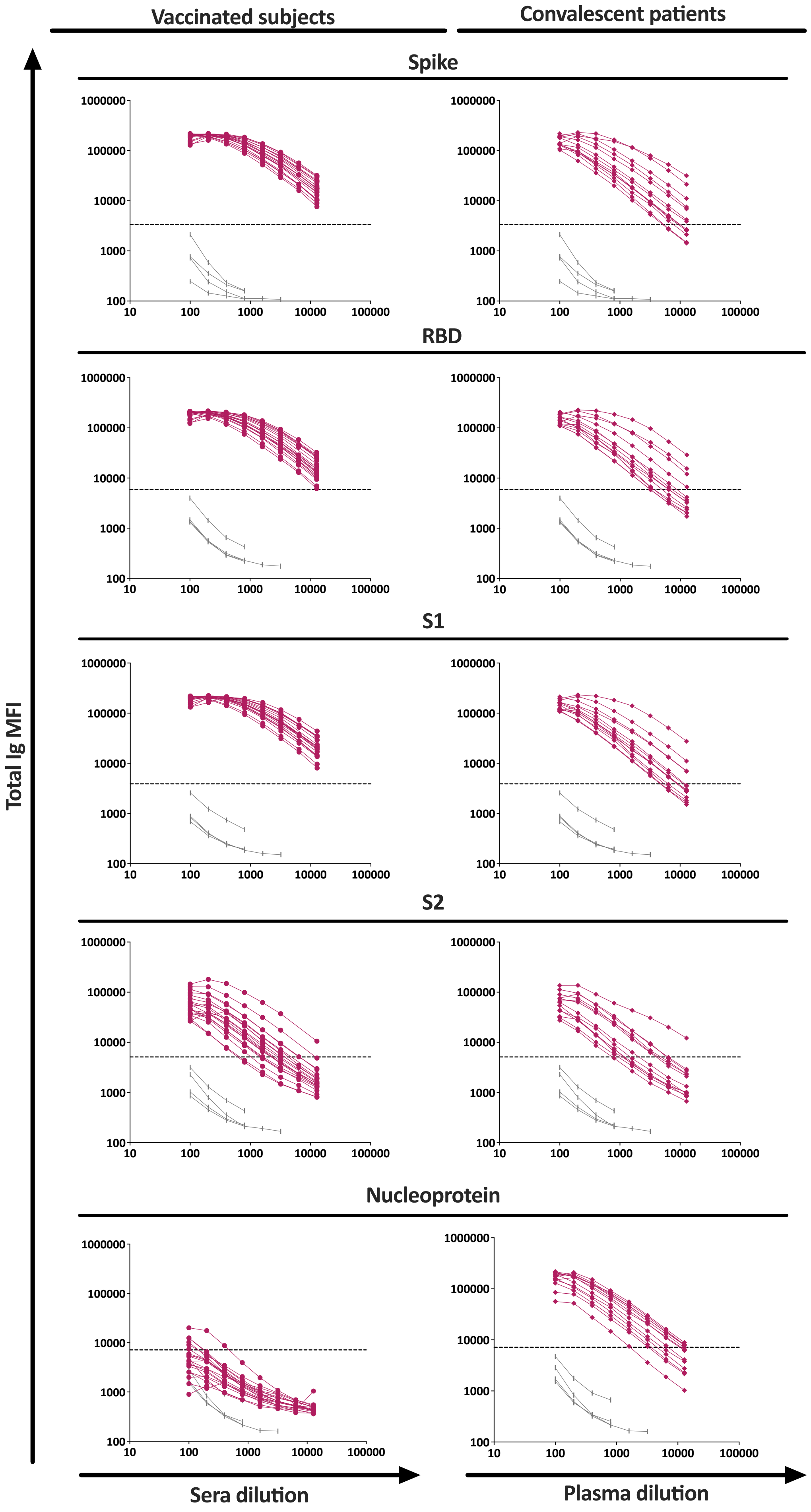

Vaccinated subjects
Convalescent patients
Spike
RBD
S1
S2
Nucleoprotein
Total Ig MFI
Sera dilution
Plasma dilution
1000000
100000
10000
1000
100
10
100000

**Supplemental Figure 5.** Titration curves are shown for total Ig against spike, RBD, S1, S2 and nucleoprotein in sera and plasma specimens from 20 vaccinated subjects (left panels), 13 convalescent COVID-19 patients (right panels) and four COVID-19-negative subjects (gray). Specimens were diluted at 4-fold dilutions from 1:100 to 1:12,800. The dotted lines indicated the cut-off values calculated as mean + 3SD of 1:100 diluted plasma of the four COVID-19-negative specimens. Data were generated using the multiplex bead antibody binding assay and mean fluorescent intensity (MFI) values were plotted.

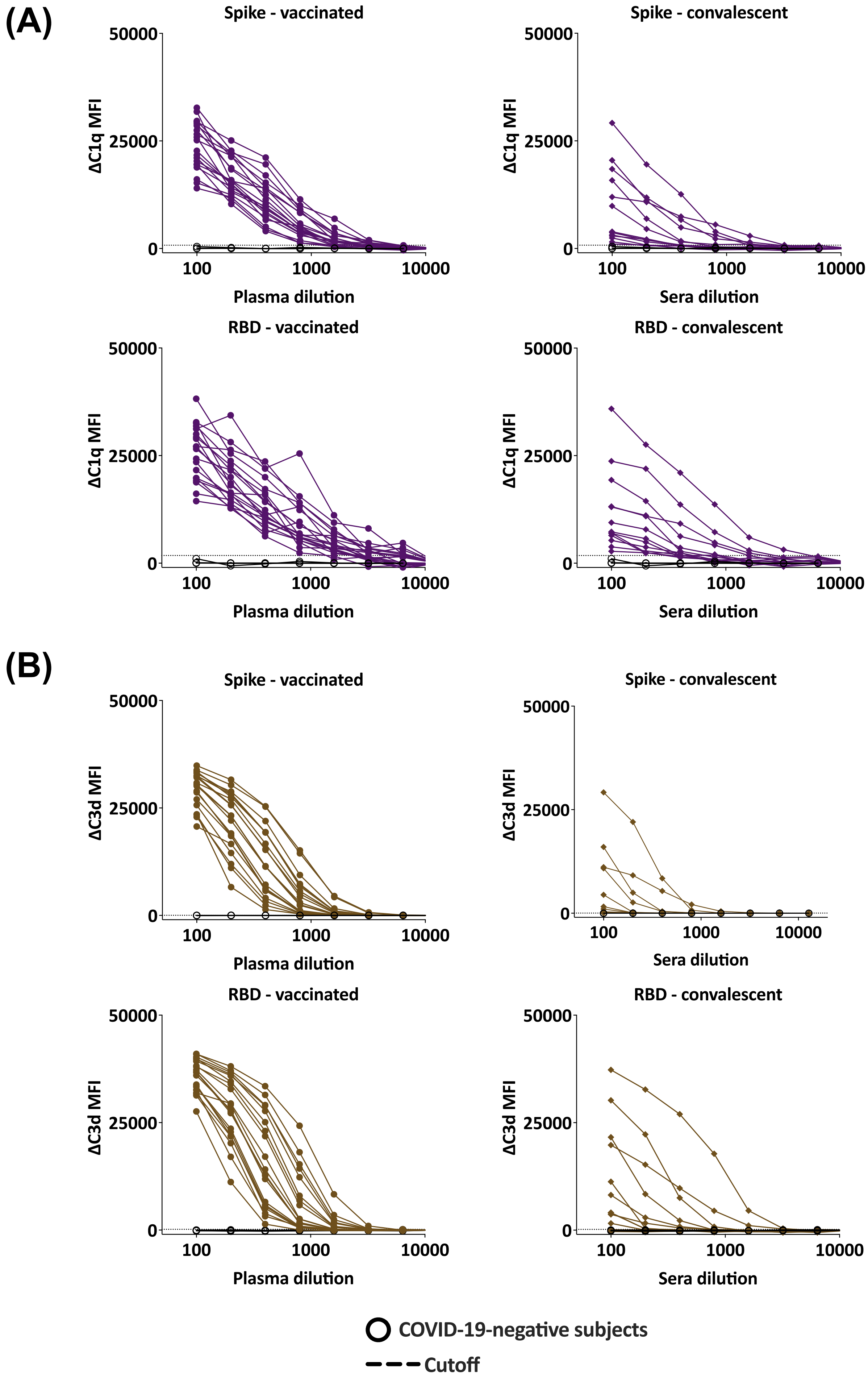
(A)
Spike - vaccinated
Spike - convalescent
RBD - vaccinated
RBD - convalescent
ΔC1q MFI
Plasma dilution
Sera dilution
(B)
Spike - vaccinated
Spike - convalescent
RBD - vaccinated
RBD - convalescent
ΔC3d MFI
Plasma dilution
Sera dilution
COVID-19-negative subjects
Cutoff

**Supplemental Figure 6.** (A-B) C1q or C3d binding to spike- or RBD-specific Abs in plasma specimens from 20 vaccinated individuals (left) and 13 convalescent COVID-19 patients (right) and four COVID-19-negative controls. Specimens were diluted at 2-fold dilutions from 1:100 to 1:6,400 or 12,800. The dotted line represents the 100x dilution cut-off calculated as mean of the four control specimens + 3SD. ΔC1q (A) and ΔC3d (B) MFI values were calculated by subtracting background MFI from each assay.
