## Supplemental Tables 1 and 2 for "Detection of Antibody Responses against SARS-CoV-2 in Plasma and Saliva from Vaccinated and Infected Individuals"

**Supplemental Table 1**

**(A)**

| Vaccinated subjects | | | | |
|---|---|---|---|---|
| **ID** | **Age** | **Sex** | **Days post-vaccination** | **Vaccine** |
| **RN#1-6** | 55-59 | F | 29 | Pfizer |
| **RN#4-2** | 60-64 | F | 23 | Pfizer |
| **RV#1** | 60-64 | M | 22 | Moderna |
| **RV#2** | 60-64 | M | 20 | Moderna |
| **RV#3** | 65-69 | M | 15 | Moderna |
| **RV#4** | 50-54 | F | 15 | Moderna |
| **RV#5** | 50-54 | M | 37 | Moderna |

**(B)**

| Convalescent COVID-19 patients | | | | |
|---|---|---|---|---|
| **ID** | **Age** | **Sex** | **Days post symptom onset** | **Disease severity** |
| **RP#2-2** | 50-54 | F | >189 | Asymptomatic |
| **RP#3-2** | 55-59 | M | 226 | Ambulatory |
| **RP#4-3** | 60-64 | M | 210 | Hospitalized |
| **RP#5-3** | 60-64 | M | 225 | Hospitalized |
| **RP#7-2** | 40-44 | F | 243 | Ambulatory |
| **RP#12** | 25-29 | M | 256 | Ambulatory |
| **RP#13** | 25-29 | F | 246 | Ambulatory |

**Supplemental Table 2**

(A)

| Vaccinated subjects | | | | |
|---|---|---|---|---|
| ID | Age | Sex | Days post-vaccination | Vaccine |
| **70771** | 35-39 | M | 27 | Pfizer |
| **39325** | 30-34 | F | 25 | Pfizer |
| **54133** | 60-64 | M | 28 | Pfizer |
| **60982** | 35-39 | M | 25 | Pfizer |
| **66685** | 30-34 | F | 29 | Pfizer |
| **83690** | 30-34 | F | 26 | Pfizer |
| **31471** | 30-34 | F | 30 | Pfizer |
| **85708** | 30-34 | M | 24 | Moderna |
| **85971** | 60-64 | F | 30 | Pfizer |
| **46611** | 30-34 | M | 30 | Pfizer |
| **82596** | 35-39 | M | 31 | Pfizer |
| **82469** | >=65 | M | 30 | Pfizer |
| **97478** | 35-39 | F | 28 | Moderna |
| **61521** | 45-49 | M | 27 | Moderna |
| **19492** | 30-34 | F | 27 | Pfizer |
| **53518** | 30-34 | F | 28 | Moderna |
| **61701** | 30-34 | F | 29 | Pfizer |
| **11150** | 60-64 | F | 28 | Moderna |
| **53676** | 25-29 | F | 30 | Moderna |
| **65670** | 40-44 | F | 29 | Pfizer |

(B)

| Convalescent COVID-19 patients | | | | |
|---|---|---|---|---|
| ID | Age | Sex | Days post symptom onset | Disease severity |
| **CVAP1** | 40-44 | M | 177 | moderate |
| **CVAP3** | 70-74 | M | 189 | severe |
| **CVAP4** | 60-64 | M | 172 | mild |
| **CVAP13** | 50-54 | M | 206 | mild |
| **CVAP14** | 55-59 | M | 207 | mild |
| **CVAP20** | 65-69 | M | 218 | mild |
| **CVAP23** | 70-74 | M | 229 | moderate |
| **CVAP24** | 75-79 | M | 216 | none |
| **CVAP31** | 65-69 | M | 214 | mild |
| **CVAP32** | 55-59 | M | 184 | mild |
| **CVAP38** | 75-79 | M | 205 | mild |
| **CVAP40** | 40-44 | M | 199 | severe |
| **CVAP41** | 55-59 | M | 134 | mild |
